# Changes in non-oscillatory features of the cortical sensorimotor rhythm in Parkinson’s disease across age

**DOI:** 10.1101/2021.06.27.21259592

**Authors:** Mikkel C. Vinding, Allison Eriksson, Cassia Man Ting Low, Josefine Waldthaler, Daniel Ferreira, Martin Ingvar, Per Svenningsson, Daniel Lundqvist

**Author notes:** **Corresponding author** Mikkel C. Vinding, Department of Clinical Neuroscience, Karolinska Institutet, Nobels väg 9, D2, 171 77 Stockholm, Sweden.

## Abstract

Parkinson’s disease (PD) is associated with changes in neural activity in the sensorimotor alpha and beta bands. Using magnetoencephalography (MEG), we investigated the role of spontaneous neuronal activity within the somatosensory cortex in a large cohort of early-to mid-stage PD patients (N = 78) and age- and sex matched healthy controls (N = 60) using source reconstructed resting-state MEG. We quantified features of the time series data in terms of oscillatory alpha power, beta power, and 1/f broadband characteristics using power spectral density, and also characterised transient beta burst events in the time-domain signals. We examined the relationship between these signal features and the patients’ disease state, symptom severity, age, sex, and cortical thickness.

PD patients and healthy controls differed on PSD broadband characteristics, with PD patients showing a steeper 1/f exponential slope and higher 1/f offset. PD patients further showed a steeper age-related decrease in the burst rate. Out of all the signal features of the sensorimotor activity, only burst rate was associated with increased severity of bradykinesia. Our study shows that general non-oscillatory features (broadband PSD slope and offset) of the sensorimotor signals are related to disease state and *oscillatory burst rate* scales with symptom severity in PD.

## 1. Introduction

Parkinson’s disease (PD) is a common neurodegenerative disease characterised by a gradual accumulation of Lewy bodies and death of dopaminergic neurons.^1,2^ The Lewy body pathology of PD begins long before the manifestation of motor symptoms. Accumulation of Lewy bodies is initially found in the olfactory bulb and brain stem and then spreads to the substantia nigra pars compacta, followed by several brain regions, including the basal ganglia and the neocortex.^3^ The progressive structural and neurochemical changes in PD are accompanied by widespread functional changes in neuronal activity, which in turn lead to worsening clinical signs and symptoms such as tremor, rigidity, and bradykinesia and co-occurring non-motor symptoms like sleep disorders, depression, fatigue, and cognitive deficits.^1^

The changes in brain function in PD are particularly prominent in the oscillatory activity of neurons.^4^ In PD, spontaneous oscillatory beta band (13–30 Hz) activity in the sub-thalamic nucleus (STN) exhibits a systematic disease-related increase in synchronicity that is related to the dopamine level^5–8^, and correlates with the severity of bradykinesia and rigidity symptoms.^9,10^ Changes in the beta band extend beyond the STN through the basal ganglia-thalamic cortical sensorimotor network. The cortical manifestation of the disease-related changes in the sensorimotor network can be measured non-invasively from the cortex, using electro- or magnetoencephalography (EEG/MEG). Such non-invasive neural recordings can potentially provide easily available prospective biomarkers of disease or symptom-related neural changes in PD. Increased oscillatory beta-band activity in the sensorimotor cortex has been linked to increased symptom severity, such as rigidity and bradykinesia.^11,12^ The role of dopamine on the cortical beta band is, however, still unclear. There is no consensus on how dopaminergic medication affects cortical beta-band power, with some studies reporting no effects^11,13–15^ and others an increase in beta-band power.^16–18^ Deep brain stimulation of the STN in PD patients has been shown to lead to a decrease in the power of spontaneous activity in the cortical sensorimotor beta and alpha (8-12Hz) bands^19,20^ (but see also^16,21^).

Importantly, there is evidence that the beta-band changes are not in the same direction across the different stages of PD. For example, there are reports of increased cortical beta-band power in the early stages of PD^22^, whereas the later stages are associated with decreased beta-band power.^23^ Further, the beta-band power is not the *only* feature of the sensorimotor rhythms that is altered in PD. Several studies have found a shift in the beta-band centre frequency (the frequency at which the power spectrum density peaks in the beta-band) towards a lower frequency in PD patients compared to healthy controls.^24–26^ The shift towards lower beta-band centre frequency is more pronounced in PD patients with dementia^27–30^ and correlates with reduced cognitive ability.^26,31^ Notably, the centre frequency shift is detectable already in the early stages of PD^25^ and does not seem to be affected by dopaminergic medication.^32^ The changes in beta-band power and centre frequency in PD could indicate that different features of the oscillatory beta-band activity reflect different underlying neural functions expressed in the measured sensorimotor signals. Changes in beta-band power could be functionally related to sensorimotor disturbances, and changes in centre frequency could be related to cognitive function.

The characteristics of neuronal oscillatory activity may hold additional information of disease-related changes in PD.^33^ Both beta-band power and centre frequency reflect a quantification of power spectral density (PSD). While these features can provide valuable information about disease-related changes in PD, the PSD quantification of a neural time series provides a static summary of the oscillatory activity across the entire time series. PSD does not account for inherent dynamics in this activity or changes in the time series on shorter time scales—as is prevalent in neural time series. The beta-band exhibits a great degree of variation over time and contains characteristic high-amplitude “bursts” that last about 50-200 ms, both in the cortical and sub-cortical beta-bands.^34–37^ Functionally, the transient bursts appear to play a pivotal role in sensorimotor processing through the basal ganglia-thalamic-cortical network. For instance, the presence of a beta burst in the sensorimotor cortex close to a tactile stimulation decreased the likelihood of tactile detection^38^, and the rate of beta bursts is shown to decrease in the time leading up to a movement both in STN^39–41^ and in the sensorimotor cortex.^42,43^

In PD, quantification of beta-band burst activity from recordings in the STN has shown that beta-burst rate and duration are reduced by dopaminergic medication^44,45^ and deep brain stimulation.^37^ Furthermore, PD patients exhibit a decrease in the rate of beta burst at the cortical level compared to healthy controls.^14^ This decrease in beta burst rate is inversely related with increased severity of motor symptoms;^46^ particulary bradykinesia and postural-kinetic tremor symptoms, but there is not evidence pointing to an effect of dopaminergic medication on cortical bursting properties.^14^ Notably, the burst rate showed a higher sensitivity than PSD beta power for discriminating PD patients from healthy controls, demonstrating that the choice of method for analysing beta-band features influences the sensitivity of subsequent analyses. This is further complicated by the fact that in addition to disease-related changes, these features likely differ with age,^43,47^ and the fact that most studies on oscillatory changes in PD come from studies with small sizes.^48^ The central challenge is quantifying the measured neural signals to extract the disease’s relevant features from the signals, be it the spectral power, centre frequencies, or burst-like features.

In the current study, we aimed to compare how different oscillatory features of cortical sensorimotor activity change in PD to elucidate what oscillatory features in the neural time-series differ between PD patients and healthy controls and how these features are associated with different motor symptoms in PD. We extracted the sensorimotor neural resting-state activity from source reconstructed resting-state MEG signals in the sensorimotor cortex (Figure 1) and quantified the time-series in terms of the PSD in the canonical mu-band (8-30 Hz).^49,50^ In addition to the band-specific analysis, we compared the 1/f broadband characteristics of the PSD.^51,52^ Finally, we compared features of the sensorimotor rhythm in terms of time-domain analysis of spontaneous transient bursts.^14,38^ We tested the hypotheses of altered functional changes in PD by analysing how these features differed between PD patients and healthy controls and further investigated the interactions with age and sex. As ageing is associated with structural and functional changes in the sensorimotor cortex^53,54^, we investigated if the potential changes in sensorimotor activity in PD differed across age. Since both healthy ageing and PD disease progression are linked to thinning of the cortex^55,56^, we further included thickness of the sensorimotor cortex in the analysis as a potential mediating factor on the sensorimotor activity that potentially also interacts with disease state.

**Figure 1:**
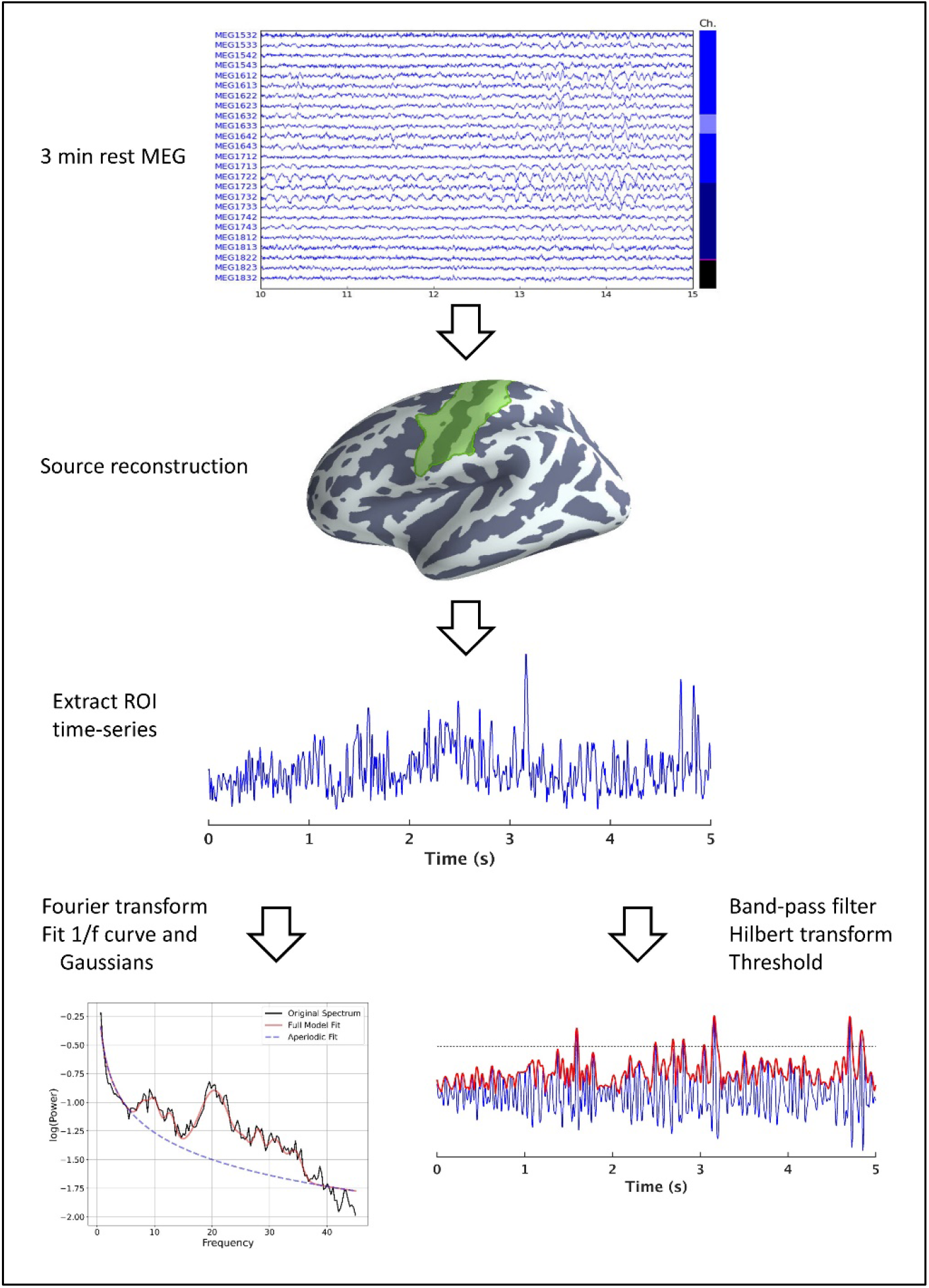
Overview of the data processing pipeline. Three minutes raw resting-state MEG data was obtained from each participant. The signals were projected through a minimum-norm source reconstruction to extract the activity in the sensorimotor cortex. We did a frequency decomposition of the source reconstructed signal to calculate the PSD to which a 1/f and Gaussian curve were fitted to extract the PSD features (Table 2). In addition, we quantified sensorimotor bursts in the signal time series in the sensorimotor ROI by thresholding the envelope of the band-pass filtered (8-30 Hz) signal to the mu-beta frequency range.

The central hypothesis was that there would be differences between healthy controls and PD in features of the sensorimotor signals, but also that different features may be related to different functional changes. We hypothesised that individual oscillatory features would reflect different underlying neural functions in the sensorimotor system and thereby show different relationships to the clinical manifestations of specific motor symptoms in PD. We tested this hypothesis in two steps: first, examining the inter-relationship between all different measures, and subsequently, examining what feature—or combination of features—best explained the variation in severity within each motor symptom.

## 2 Results

To enable a sensitive assessment of disease-related oscillatory changes in PD, we aimed for a relatively large cohort compared to other functional neuroimaging studies of PD patients (N=78) and healthy controls (N=60), balanced across gender and age, and with gender- and age-matched groups (Table 1).

**Table 1:** Group-level summary of the participants included in the analysis. Mean (standard deviation).

| Measure | Parkinson’s patients | Healthy controls | Statistics |
| --- | --- | --- | --- |
| *N* | 78 | 60 |  |
| *Sex (female/male)* | 29/49 | 27/33 | $\chi^2 = 0.57$, $p = 0.45$ |
| *Age* | 65.6 (9.5) | 63.93 (8.4) | Welsh $t(138.0) = 1.08$, $p = 0.28$ |
| *Disease duration* | 4.4 (3.7) years | - | - |
| *LEDD* | 548 (273) mg | - | - |
| *MDS-UPDRS-III* | 18.9 (10.8) | - | - |
| *MoCA* | 26.1 (2.8) | 26.2 (2.1) | Welsh $t(136) = 0.10$, $p = 0.92$ |

For the first analysis, we investigated how features in the resting-state activity from the sensorimotor area quantified by features of the PSD and burst characteristics (see Table 2) differed as a function of the predictors *group* (PD patients/healthy controls; Table 1), *age, sex*, and *cortical thickness* as well as the interaction between the predictors.

**Table 2:**
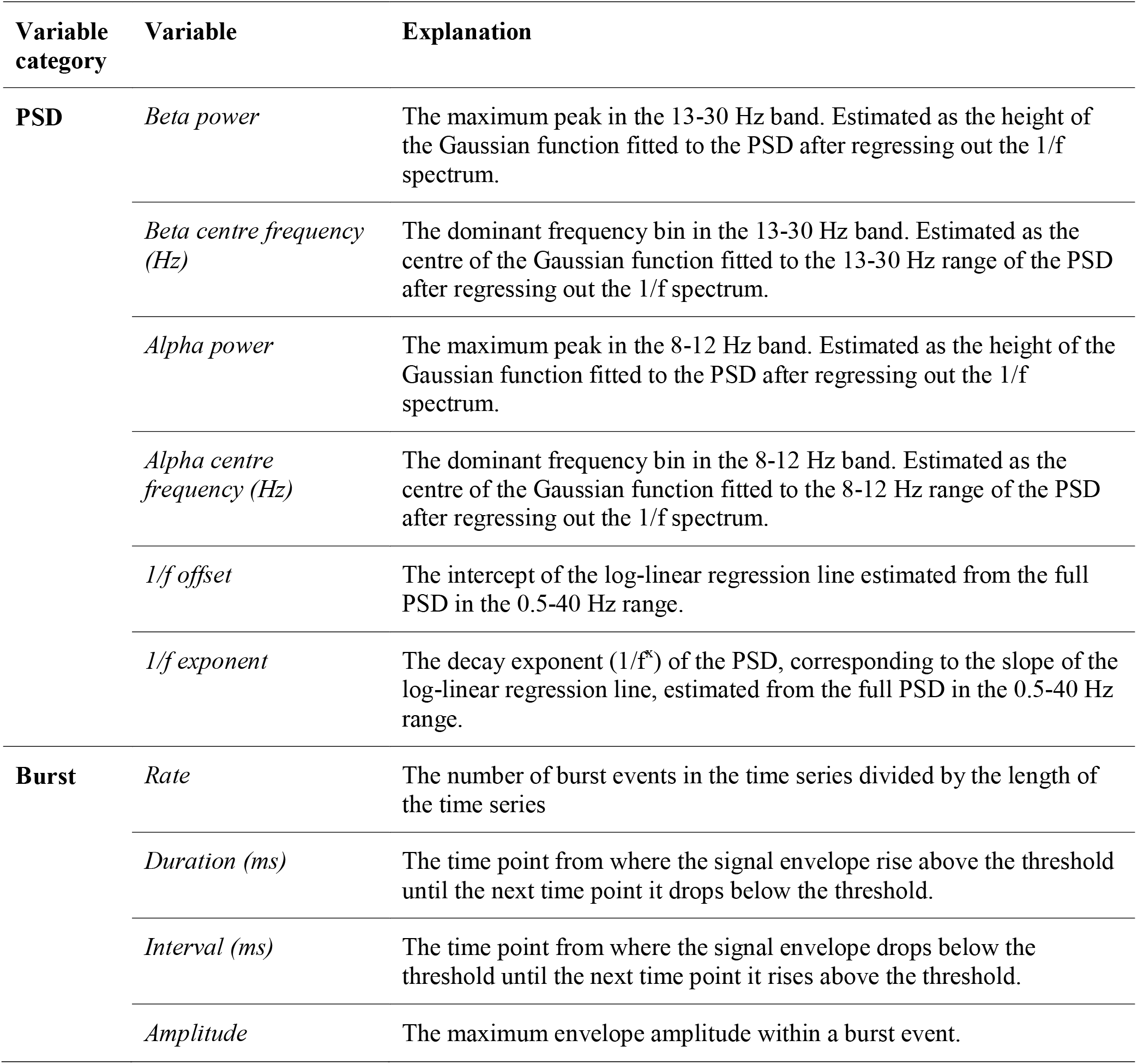
Summary explanations of the main outcome variables in the analysis

### 2.1 PSD features

The PSD suggests an apparent group difference between PD patients and healthy controls in the alpha and beta bands (Figure 2). However, analysing the oscillatory components of PSD by first adjusting for the broadband characteristic of the PSD^51^ removed the apparent group difference in the mu- and beta band. Bayesian model comparison was used to test which predictors explained the variation in the PSD (Bayes Factors (BF) > 3 taken as cutoff for substantial evidence for an effect of a given predictor^57^). The model comparison showed evidence for group differences on the 1/f offset (BF = 37.77) and 1/f exponent (BF = 5.92). There were no substantial evidence of an effect beyond the threshold for substantial evidence on any other PSD features. However, it might be worth noting that there was anecdotal evidence of an effect of cortical thickness on 1/f exponent (BF = 1.73), an interaction effect between age and group on alpha centre frequency (BF = 1.43), and only minute evidence for a group difference on beta power (BF = 1.28). The coefficients of regression models analysing the effect of the predictors on PSD for all outcome measures are presented in Figure 3.

**Figure 2:**
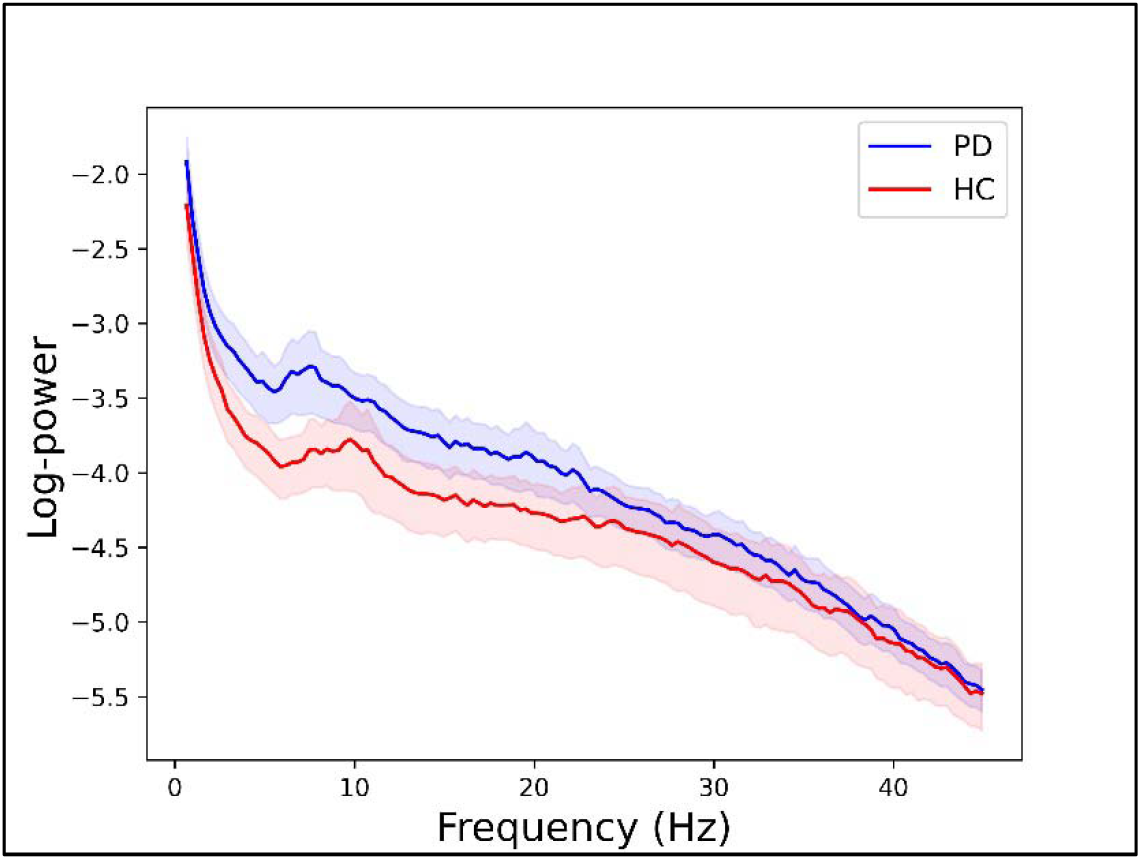
Group-level PSD. Grand average PSD (mean+standard error) for the PD group (blue) and healthy control group (red).

**Figure 3:**
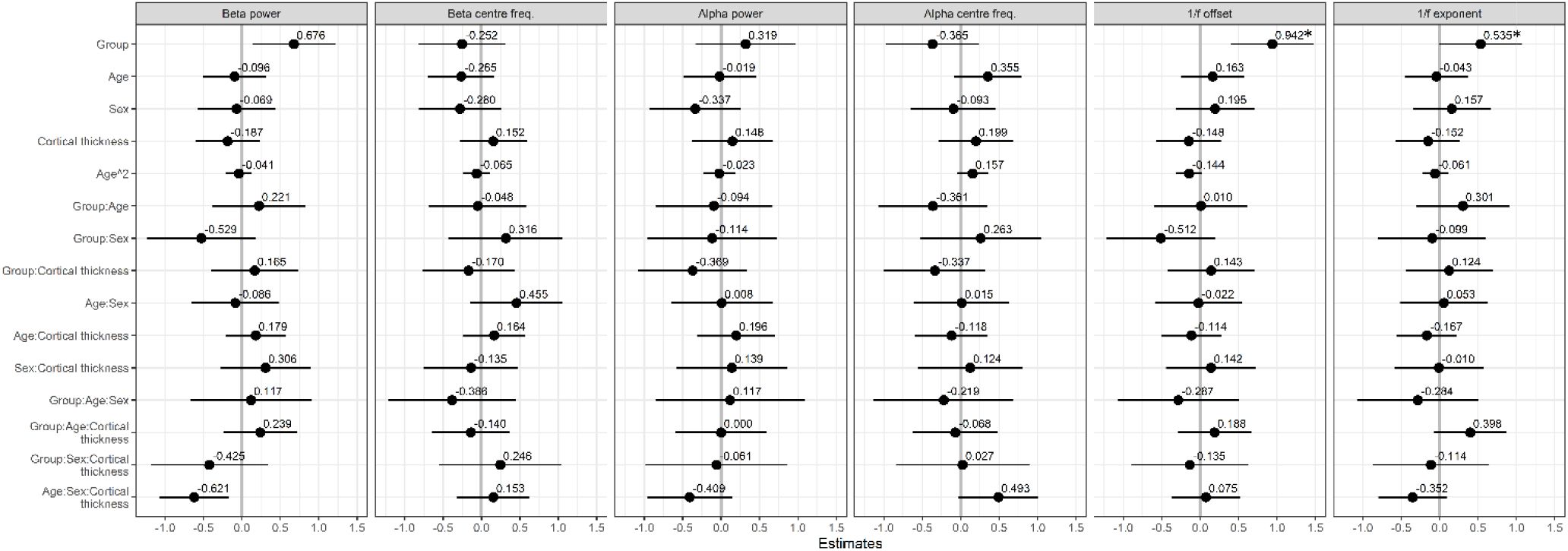
Regression analysis of PSD features. Standardized regression coefficients for the analyses of sensorimotor PSD features as a function of group, age, sex, cortical thickness and the interaction between these factors. * indicate factors with Bayes Factor > 3 in the model comparison.

Analysing the PSD by regressing out the 1/f contribution to the spectrum, the difference between PD patients and healthy controls is not so much in the canonical beta- and alpha bands but manifests in the broadband characteristics of the signal. The 1/f intercept was 23.5% [CI: 11.8:33.7%] higher for PD patients than healthy controls, and PD patients had on average 11.9% [CI: -30.4:3.8%] steeper 1/f exponential slope compared to healthy controls.

### 2.2 Burst features

The view of sensorimotor oscillatory activity has recently changed from a steady oscillating signal to viewing the activity in the sensorimotor bands occurring in short bursts. We compared features of the sensorimotor rhythm in terms of time-domain analysis of spontaneous transient bursts in the time series. Model comparison to test which predictors explained the variation in the burst features gave evidence for an interaction effect between group and age on the burst rate (BF = 33.57) and a main effect of sex on the burst rate (BF = 4.34). Model coefficients are displayed in Figure 4.

**Figure 4:**
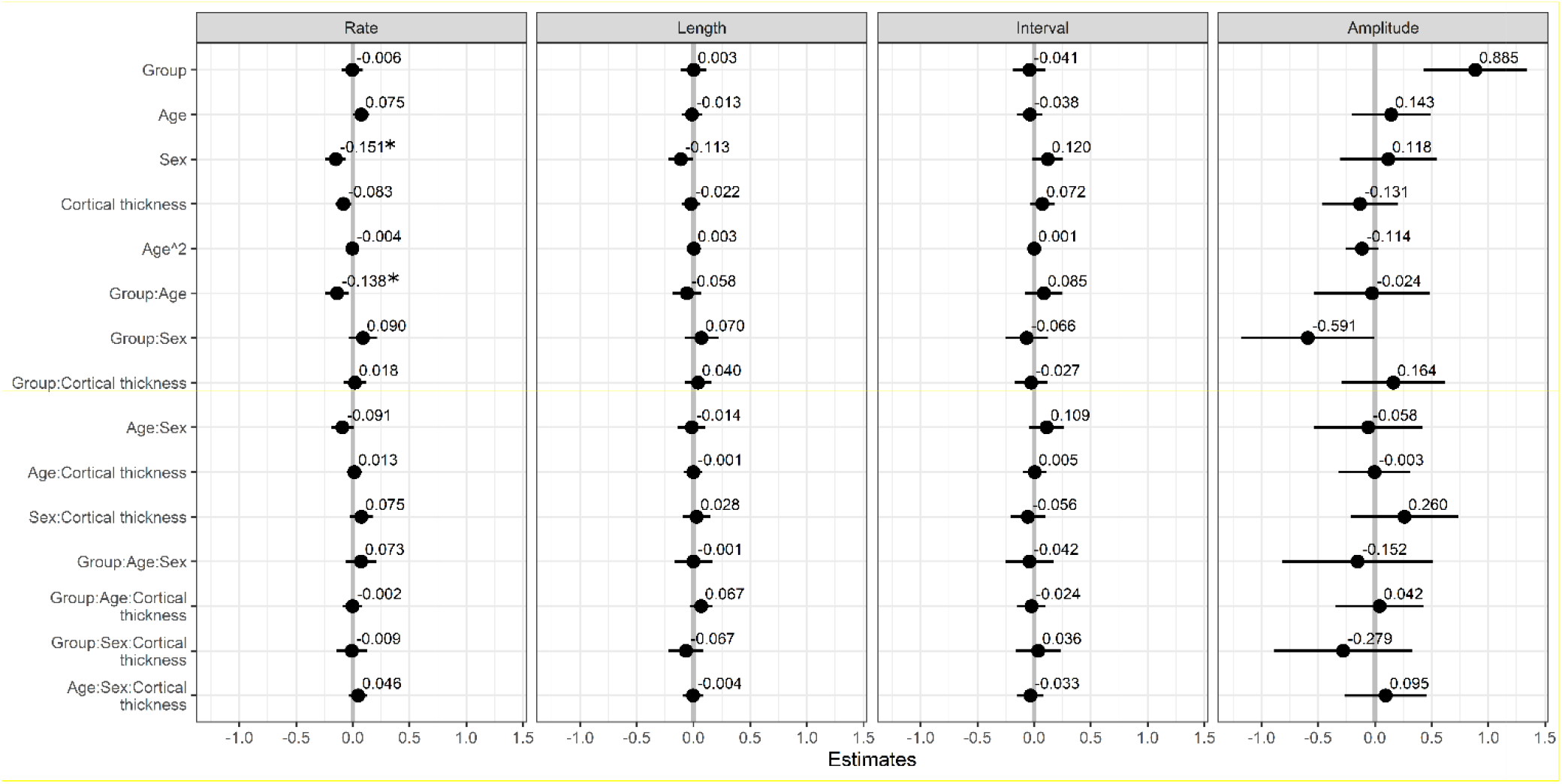
Regression analysis of burst features. Standardized regression coefficients for the analyses of burst features as a function of group, age, sex, cortical thickness and the interaction between these factors. * indicate factors with Bayes Factor > 3 in the model comparison.

The age-related effects from the model amount to a change in burst rate of -0.7% [CI: -1.6:0.2] per year for female PD patients and -1.7% [CI: -3.1:-0.3] change in burst rate per year for male PD patients, whereas female controls had a relative increase in burst rate of 0.8% [CI: 0.1:1.7] per year and male controls had a stable trend of -0.2% [CI: -1.0:0.6] change per year.

No other predictors showed evidence of an effect beyond the threshold for substantial evidence on neither burst length, the interval between bursts, nor burst amplitude. There was anecdotal evidence (i.e. 1/3 < BF < 3) for a group difference in burst amplitude (BF = 2.08) as well as an interaction effect of age and cortical thickness on the burst rate (BF = 1.43), and an interaction between sex and cortical thickness on the burst rate (BF = 1.86).

### 2.3 Clinical symptoms and oscillatory features

In the second analysis, we tested for associations between motor symptoms and the features of the sensorimotor signal in the PD group. All sensorimotor signal features listed in Table 2 were used as predictors in a multiple regression analysis that further included age, sex, and cortical thickness to regress out the contribution hereof. The standardised regression coefficients of each predictor variable on the motor symptoms measured with MDS-UPDRS-III^58^ are presented in Figure 6.

**Figure 5:**
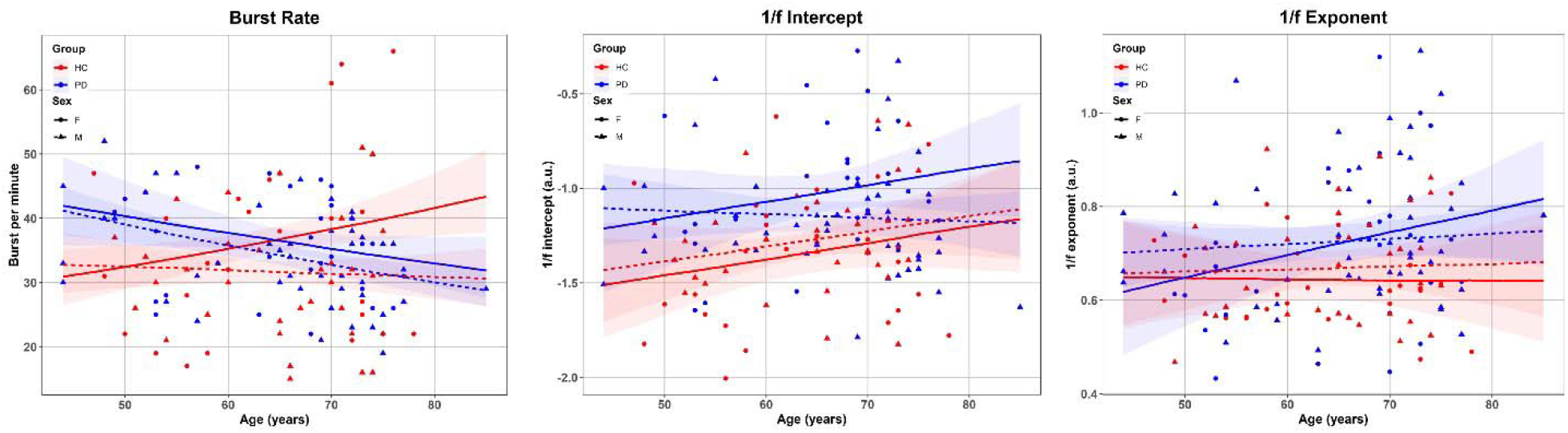
Age-related differences in sensorimotor signal features. Scatterplots of the individual measures and model predictions over age for (A) burst rate, (B) PSD broadband 1/f intercept, and (C) PSD broadband 1/f exponent, split between PD patients (blue) and healthy controls (red), and female (solid) and male (dashed).

**Figure 6:**
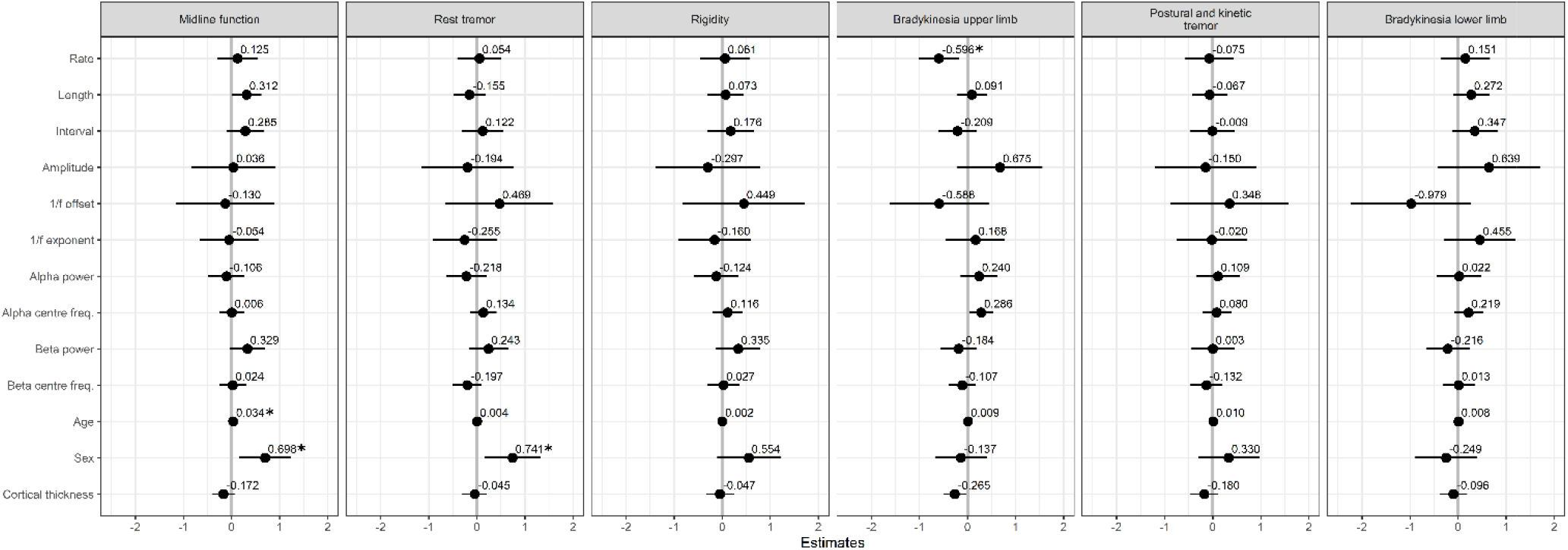
Regression analysis of motor symptoms. Standardized regression coefficients for the regression analyses of sensorimotor signal features on clinical motor symptom ratings in PD. * indicate factors with Bayes Factor > 3 in the model comparison.

Model comparison of multiple regression models showed evidence that burst rate was negatively associated with upper limbs bradykinesia (BF = 12.70). The negative direction of the effect of burst rate means that reduced burst rate was associated with increased severity of bradykinesia. The analysis yielded no substantial evidence for effects of other sensorimotor signal features on symptom ratings for axial symptoms, rest tremor, rigidity, rest tremor, postural/kinetic tremor, nor lower limb bradykinesia.

The analysis yielded substantial evidence for an effect of age on axial symptoms (BF = 5.79) and evidence for a difference between male and female patients on axial symptoms (BF = 7.56) and rest tremor (BF = 6.06). There were anecdotal evidence for an effect of alpha centre frequency on upper limbs bradykinesia (BF = 2.63), an effect of burst length on axial symptoms (BF = 1.49), as well as anecdotal evidence for an effect of cortical thickness on upper limbs bradykinesia (BF = 2.63).

## 3 Discussion

This study aimed to explore how different features of cortical somatosensory oscillatory activity at rest differed between PD patients and healthy controls across age and gender and how these features relate to motor symptoms in PD. The analysis of spontaneous sensorimotor bursts showed an increased age-related reduction in PD patients compared to healthy controls. Notably, our current results show that the reduced burst rate in PD is not a static group-level difference but interacts with age with a steeper age-related reduction in burst rate in PD compared to healthy controls.

We hypothesised that different oscillatory features reflect distinct underlying functional neural properties and manifest as different motor symptoms in PD. The results showed that a reduction in mu-beta burst rate in PD was accompanied with an increase in bradykinesia severity, confirming previous findings from our group.^14^ This relationship was exclusive for the bursts rate, as we observed no statistically evident relationships between other burst features or PSD features and clinical motor symptoms.

### 3.1 Characterizing neural time series data

The cortical sensorimotor activity of the PD patients differed from healthy controls, but did not differ on the PSD in the canonical sensorimotor mu and beta bands. The PD patients showed a steeper broadband 1/f slope and exponent of the broadband PSD than healthy controls. The observation of a group difference in the spectral broadband 1/f characteristics of the signal adds to the growing evidence that a focus on neural activity as narrow-band steady oscillations—e.g. narrowly focusing only on the beta-band power—could potentially miss essential aspects of the neural signals for understanding mechanistic changes in disease.^59,60^ Widening the quantitative analysis of PSD rather than focusing exclusively on narrow band activity is of potential clinical value: quantifying only the peaks in the PSD to differentiate patients from controls can misrepresent the actual oscillatory response at those frequencies as the peaks are influenced by the broadband offset and 1/f exponent. Any unaccounted-for systematic differences in either PSD offset or decay exponent can lead to a false conclusion that there is a difference in the oscillatory response.^51,52^ Non-invasive measurements of changes in sensorimotor activity is—despite the often conflicting findings^48^—a potential useful method to assess disease-related changes. At the current stage of the field there is, however, a need to further bridge how features in the signals are linked to disease mechanisms.

Analysis of neural time series by frequency decomposition is a powerful tool to extract and summarise features of the signal, but the method comes with limitations in what one can infer. In the time domain, increased oscillatory power can reflect both increased burst duration and change in burst amplitude and as an expression of true sustained oscillations in the signal.^36,61^ The presence of more sustained oscillations in the sensorimotor rhythm might reflect a higher level of inhibition of sensorimotor information; as is seen in recordings from STN^5^ and, to some extent, also at the cortical level.^38,62^ However, sustained oscillations are not in contrast to the bursting properties of the sensorimotor rhythm. The neural time-series can express both a degree of sustained oscillations while also exhibiting variation in the degree of transient bursts—e.g., a signal of steady oscillation with transient high-amplitude bursts.

### 3.2 Beta bursts and beta activity

Bursting properties of the cortical sensorimotor neural activity are proposed to occur due to long-range input through the ascending thalamic-cortical connection to the cortex, leading to an increase in the local neural excitation and resulting in a burst of synchronous activity.^36^ The observed disease-related changes in spontaneous cortical bursts, in the form of a more rapid decrease in rate over age for PD patients, could reflect inhibition of these projections along the thalamic-cortical pathways caused by disturbances in the dopamine-dependent structures projecting to the cortex. Interestingly, we did not find significant group differences in burst duration in the current study—in line with previously reported findings on cortical burst in PD^14^—supporting the view that the central mechanisms of the cortical bursts are not primarily affected in PD—instead, it is the *rate* of bursts that is reduced at the cortical level. The sub-cortical beta-band activity is influenced by the activity of dopamine-responding neurons^5,6,45^. The effect of dopamine and dopaminergic medication on the cortical beta-band is likely mediated by the dopaminergig neurons projecting to the cortex that terminates in the pre-fontal cortex but also to less extend in the primary sensorimotor cortex.^63^ The differences in the cortical sensorimotor burst rate in PD might be an indirect effect of the loss of dopamine and changes in the beta band in the sub-cortical structures projecting to the sensorimotor cortex. The notion that the cortical sensorimotor activity is indirectly related to dopamine depletion in PD is further supported by findings from animals studies showing that 6-hydroxydopamine injections lead to exaggerated beta-band oscillations only after several days had passed, suggesting that oscillatory changes occurred as an indirect compensatory effect after dopamine depletion rather than a direct consequence of the depletion itself.^7^ The indirect influence of dopamine on the cortical beta-band might also explain the often weak or even absent effect of dopaminergic medication on cortical beta-band activity.^11,13–15^ The current study cannot directly address the role of dopamine on cortical oscillations since all patients in the study were tested on medication. However, a recent study found that cortical burst characteristics measured with MEG were influenced by DBS therapy in PD and normalised the bursting chracteristics during DBS to resemble the burst characteristics of healthy controls.^46^ This further suppots that cortical bursting activity is mediated by subthalamic projections.

### 3.3 Age-related differences

We explored how age-related differences in cortical sensorimotor neural activity might interact with disease-related changes in PD. Age-related effects on spontaneous sensorimotor activity are commonly dealt with by matching the age distributions of the patient group and the healthy control group— usually within a narrow age span. The analysis showed age-related differences in burst rate, with PD patients showing a more considerable reduction of burst as a function of age than healthy controls. The steeper reduction in burst rate with age in PD seems in accordance with the fact that higher age at PD onset is associated with a faster disease progression and more rapid decline in motor function^64^, though a longitudinal design is needed to confirm the relation between disease progression, reduction in burst rate, and age. We did not see a significant “slowing” of the beta PSD centre frequency between groups, as reported in several previous studies.^48^ An explanation might be that such slowing is more pronounced in PD patients with dementia^27–30^ and correlates with cognitive ability.^26^ The PD patients in the current study did not differ in their cognitive ability from the healthy controls. Furthermore, we focused on the activity in the sensorimotor cortex, whereas the slowing of alpha and beta PSD is usually found in frontal areas and globally throughout the brain.^25,28,31^ We included cortical thickness measures within the same ROI from which we extracted the functional time-series, as we hypothesised that age-related effects upon the functional measures might be mediated through the age-related structural changes in the cortex. However, despite the negative correlation between age and cortical thickness (see supplementary material), we did not find pervasive evidence that cortical thickness affected any of the functional measures.

### 3.4 Sex differences

We also included sex to explore if disease-related changes in sensorimotor oscillatory activity differed between males and females, as there are well-documented sex differences in the manifestation of PD.^1,65^ Male sex is a risk factor for developing PD, with an average incidence ratio of approximately 2:1 male-female ratio across all stages of the disease.^66^ The disease onset is on average two years earlier in males than females and differs in the initial manifestation of symptoms, with women more likely to develop tremor specific symptoms and men more likely to develop rigidity.^67^ We are not aware of any previous studies that explicitly included sex as a factor in analysing neural oscillations in PD. The regression analysis of motor symptoms showed evidence for a difference in midline function and rest tremor between male and female patients. We did not, however, find widespread sex differences in the various features of the sensorimotor activity. The analysis of the sensorimotor singal features only showed evidence for differences between males and females in the burst rate. A possible factor behind the sex differences in PD is the contribution of sex hormones on the nigrostriatal pathway and linked to the deterioration of the dopaminergic system, where testosterone levels appear to enhance dopamine loss, while estrogen has been identified as a neuroprotective agent for PD. Estrogen has been demonstrated to influence incidence levels of PD while menopause-related variations in estrogen levels are linked to variations in PD symptom severity.^68^ At the current stage, it is unclear if estrogen sex hormones influences oscillatory bursts in the sensorimotor cortex. These findings illustrate the need for further studies into sex-specific changes in neural function and how they manifest and relate to PD.

### 3.5 Limitations and conclusions

The present study quantified the neural time series from the sensorimotor cortex based on pre-defined summary measures of its PSD and burst properties. We included more PD patients and healthy controls than similar previously conducted functional studies—typically in the range of 5-30 participants.^48^ A limitation of our study for understanding the extent of changes in oscillatory sensorimotor activity is the focus on different features within a narrow ROI, which ignores other types of measurements that are potentiallyrelevant to understanding the development of PD and motor symptoms: for example, the long-range connectivity between the sensorimotor cortex and other cortical areas and the connections between the sensorimotor cortex and the basal ganglia and thalamus (though the subcortical structures are practically invisible in MEG).

Treating the activity in the sensorimotor cortex as single time series also means that we remove the sensitivity to spatial features of the signals, e.g., focal versus spatially blurred activity in one group or the other. If the oscillatory activity extends over a larger cortical surface area, that signal will also manifest as power differences in the measured signal.^61^ There are potentially other features to be uncovered, and future studies may explore how the PSD- and burst features further interact with other aspects of brain activity in the global function of the brain to fully understand the interaction between functional and structural changes in PD.

We investigated a relatively large cohort of PD patients and healthy controls (for a neuroimaging study) to make meaningful inferences about how age and sex interact with the group level difference between PD patients and healthy controls; however, a limitation is that our study is cross-sectional. We aim to follow this cohort longitudinally to estimate the development trajectories of the sensorimotor oscillatory activity in PD compared to healthy ageing.

Sensorimotor activity measured non-invasively with MEG/EEG contains rich information about the functional state of the sensorimotor system and how it changes in PD. The central challenge is quantifying the measured neural signals to extract the disease’s relevant features from the signals, be it the spectral power, centre frequencies, or burst-like features. Finding features of neural signals that can explain disease mechanism or symptoms, even if extracted along with a reduced number of dimensions, will be helpful if they provide adequate information about the disease- or symptom-state. Further characterisation of the association between features in the non-invasive brain signals and motor symptoms can potentially be a valuable tool to aid in diagnosis and treatment evaluation. Understanding how features in the neural time series are related to motor symptoms in PD will also help develop non-invasive neural stimulation that can potentially relieve motor symptoms.^37,69^

## 4 Methods

### 4.1 Participants

Eighty PD patients (age 44-85; 32 female) and 71 healthy controls (age 46–78; 46 female) participated in the study. The study was approved by the regional ethics committee (Etikprövningsnämden Stockholm, DNR 2019-00542) and followed the Declaration of Helsinki. All participants gave written informed consent before participation.

The PD patients were recruited from the Parkinson’s Outpatient Clinic, Department of Neurology, Karolinska University Hospital, Stockholm, Sweden. The healthy controls were recruited by advertising or amongst spouses of PD patients. 22 participants (18 patients, 4 healthy controls) were included from a previous study^14^ who were qualified based on the recruitment criteria of the present study and had done the same MEG and MRI procedures as in the present study. All data were reanalysed following the procedure described below.

The inclusion criteria for the PD group were a diagnosis of PD according to the United Kingdom Parkinson’s Disease Society Brain Bank Diagnostic Criteria with Hoehn and Yahr stage 1-3.^70^ Inclusion criteria for the control group were not having a diagnosis of PD, no form of movement disorder, and no history of neurological disorders, epilepsy, or psychiatric disorders.

Exclusion criteria for both groups were a diagnosis of major depression, dementia, history or presence of schizophrenia, bipolar disorder, epilepsy, or history of alcoholism or drug addiction according to the *Diagnostic and Statistical Manual of Mental Disorders*.^71^

One participant declined to do the MRI scanning, one participant had a scanner malfunction during MRI acquisition, and 11 participants had their MRI scans cancelled due to the Covid-19 pandemic and were not included in the analysis. In total, two PD patients and 11 healthy controls were excluded from the analysis. Table 1 is a summary of the participants included in the analysis.

The PD patients participated in the study while on their regular prescribed dose of medication. The levodopa equivalent daily dose (LEDD) was calculated according to Tomlinson et al.^72^ Motor symptoms in the PD group were assessed using the motor section of the Movement-Disorder Society Unified Parkinson’s Disease Rating Scale (MDS-UPDRS-III).^59^ Global cognition was assessed with the Montreal Cognitive Assessment battery (MoCA).^74^

### 4.2 MEG recordings

MEG data were recorded with a Neuromag TRIUX 306-channel MEG system, with 102 magnetometers and 102 pairs of planar gradiometers. Data were sampled at 1000 Hz with an online 0.1 Hz high-pass filter and 330 Hz low-pass filter. The MEG scanner was located inside a two-layer magnetically shielded room (Vacuumschmelze GmbH) with internal active shielding active to suppress electromagnetic artefacts. The subjects’ head position and head movements inside the MEG scanner were measured during recordings with head-position indicator coils (HPI) attached to subjects’ heads. The HPI location and additional points sampled uniformly across the subjects’ head shape were digitalised with a Polhemus Fastrak motion tracker before the measurements. Horizontal and vertical electrooculogram (EOG) and electrocardiogram (ECG) were recorded simultaneously with the MEG.

We recorded three minutes of resting-state MEG while the participants sat with their eyes closed. The participants were instructed to close their eyes and relax. The recordings began after assuring the participant sat still with their eyes closed.

### 4.3 MRI acquisition

3D T1-weighted magnetisation-prepared rapid gradient-echo (MPRAGE) sequence structural images (voxel size: 1×1×1 mm) were obtained on a GE Discovery 3.0 T MR scanner for morphological analysis and creating source spaces for MEG source reconstruction. Multi-echo “FLASH”^75^ images were obtained to create volumetric headmodels for MEG source reconstruction (see below).

### 4.4 MRI processing

The MRI images were processed with Freesurfer^76^ (v. 5.3) to get surface reconstructions of the cortical mantle. The surfaces were obtained with the automatic routine for extracting cortical surfaces in Freesurfer from the individual T1-weighted MRI.

We defined the cortical sensorimotor area by segmenting the cortical surface using the anatomical labels provided by Freesurfer automatic labelling.^77^ The analysis focused on a region of interest (ROI) consisting of the left pre- and post-central gyri and central sulcus. The pre/postcentral gyri were combined because a biomagnetic source on either sulci wall will leave a trance on the other side due to the close distance and the field spread of MEG signals. The ROI was defined for each subject based on the individual cortical reconstructions. The average cortical thickness in the ROI was estimated with Freesurfer.^78^

### 4.5 MEG pre-processing

The MEG data was processed by applying temporal signal space separation (tSSS) to suppress artefacts from outside the scanner helmet and correct head movement during the recording.^79^ The tSSS had a buffer length of 10 s and a cut-off correlation coefficient of 0.95. Movement correction was done by shifting the head position to a position based on the median of the continuous head position during the three-minute recording.

The MEG data processing and source reconstruction was done with MNE-Python^80^ in Python 3.8. First, we marked data segments containing muscle artefacts and SQUID jumps with the automatic artefact detection in MNE-Python. The data was filtered with a 48 Hz low-pass filter and 50 Hz notch filter to remove line noise. The continuous data were cut into 1.0 s epochs, and epochs with muscle artefacts or extreme values (5000 fT for magnetometers and 4000 fT/cm for gradiometers) were rejected. Between 0-65 % (median: 6.0 %) of data was rejected resulting in 63.0-180 s (median: 174.0 s) of useful MEG data per participant. The remaining data length was not significantly different between groups (Wilcoxon rank sum test, p = 0.98). We then performed an independent component analysis (ICA) using the *fastica* algorithm^81^ to identify artefacts from blinks and heartbeats. Components showing correlation with the EOG and ECG were removed from the raw data. Between 0-5 (median 3) components were removed per participant. The number of removed ICA components was not significantly different between groups (Wilcoxon rank sum test, p = 0.71).

We then applied source reconstruction using noise weighted minimum-norm estimates.^82^ The noise covariance matrix was estimated from two minutes of empty room MEG data recorded before each session. The source space consisted of 5124 evenly spaced points sampled across the white matter surfaces. The inner skull boundary was estimated from the multi-echo MRI to create a single shell volume conductor model. The time series from the sensorimotor ROI (see Figure. 1) was extracted from the estimated source time series by singular value decomposition of all source points within the ROI.

### 4.6 Power spectral analysis

We analysed the spectral properties of the sensorimotor activity by calculating the PSD from 0.5 to 40 Hz across the entire cleaned ROI time series using Welch’s method by segmenting the continuous data into 3.072 s epochs with 50% overlap and averaging the PSD across the segments.

Since the narrow-band beta power in the PSD is dependent on the broader features of the broadband spectrum, we further analysed the 1/f broadband characteristic of the sensorimotor activity as this could play a role in the functional properties of the beta-band and has been shown to differ between healthy control and PD patients.^14^ We used the *fitting oscillations & one over f* (FOOOF) toolbox^51^ to analyse the 1/f broadband characteristic of the PSD (intercept and exponent) and the oscillatory peaks in the canonically defined beta band (13-30 Hz) and alpha band (8-12 Hz). A log-linear regression is fitted to the PSD and subtracted before fitting Gaussian functions to the peaks in the PSD. The midpoint of the Gaussian function fittied to a given frequency band corresponds to the peak frequency in that frequency band and the height represents the signal power. A new log-linear function is fitted to the PSD after subtracting the Gaussian function to estimate the 1/f characteristic.

All participants showed a discernible beta peak in the PSD. Nine PD patients and nine healthy controls did not show a peak in the PSD alpha band (no difference between groups, χ^2^(1) = 0.12; p = 0.73).

### 4.7 Burst analysis

To calculate the burst properties of the sensorimotor activity in the time domain, we band-pass filtered the time-series with an 8-30 Hz band-pass filter using FieldTrip^83^ in MATLAB (R2016b; MathWorks Inc.) and calculated the Hilbert envelope of the signal. The burst threshold was defined as two times the median of the signal. The burst onset was defined as the time-point where the signal first reached half the max amplitude of the burst and ended at the time-point where the signal again dropped below half the max amplitude of the burst. The *burst amplitude* was defined as the maximum value of the burst. The burst *duration* was defined as the time from burst onset to burst end. The *burst interval* was defined as the time from the end of a burst to the time-point where the next burst began.

### 4.8 Statistics

#### 4.8.1 Analysis of sensorimotor rhythm features

The main analyses tested the effect of *group* (PD patients/healthy controls), *age, sex*, and *ROI cortical thickness* on the features listed in Table 2. For the PSD features, we modelled the outcomes as a linear function of group (PD patients/healthy controls), age, age squared, sex, and cortical thickness with linear regression in *R* (v. 4.0.2).^84^ The regression models were fitted to the data for each participant with all factors and up to their three-way interactions between the four predictors. Gaussian regression models were estimated for each feature, except for the burst rate (burst per minute), which was modelled with Poisson regression using the same predictor variables. Before fitting the regression models, burst duration, burst interval, and burst amplitude were log-transformed.

Significance testing was done by removing one predictor from the model and comparing the variance explained between the full model and the model with a predictor removed. Hypothesis testing was done by computing the Bayes factor (BF) for the model with a given predictor (H1) versus the model without the predictor (H0) using the BIC approximation.^85^ The BF tells how much more likely the observed data is under the alternative model (H1) versus the null model (H0). The Bayesian model comparison, therefore, avoids the multiple comparison problem of frequentist hypothesis testing (theoretical likelihood of the hypothesis given the observed data). The model comparison approach furthermore circumvents issues with the interpretation of p-values of individual regression coefficients due to internal correlation between predictor variables. Following the conventional interpretation of BFs, we used BF > 3 as a cut-off between anecdotal evidence and substantial effects.^57^ The BFs for all comparisons are presented in Supplementary Table S3.

#### 4.8.2 Clinical scores and sensorimotor oscillatory features

The MDS-UPDRS-III scores were divided into subscales based on symptoms: *midline function, rest tremor, rigidity, upper-body bradykinesia, postural and kinetic tremor*, and *lower limb bradykinesia*; according to Goetz et al.^86^, with the exception that left- and right-side upper-body bradykinesia were combined into a single factor.

Each symptom score was analysed by multiple regression and modelled as a function of the burst rate, median burst duration, median bursts interval, median burst amplitude, PSD 1/f intercept, PSD 1/f exponent, PSD beta power, PSD beta centre frequency, PSD alpha power, and PSD alpha centre frequency for each PD patient. The models further included the age, sex, and cortical thickness to regress out the contribution hereof and estimate the relative effect size of each signal feature. All symptom ratings and continuous predictor variables, except age, were z-transformed to get the standardised effect size. Significance testing was done by removing one predictor from the model and calculating the BF between the full model (H1) and the model without the predictor (H0) using the BIC approximation for BFs. The BFs for all comparisons are presented in Supplementary Table S4.

### 4.9 Data Availability

The full dataset cannot be made publicly available, as the ethical permits for the study does not allow for open data sharing. Parts of the data used in this analysis will be made available as part of an online data repository (Vinding, et al. *in prep*). The scripts used to process the data and run the analysis presented in the paper are available at: https://github.com/natmegsweden/PD_beta_bursts2.

## Data Availability

Data will be made available fall 2022.

## Authorship contribution statement

**Mikkel C. Vinding**: conceptualization, methodology, software, validation, formal analysis, investigation, data curation, writing - original draft, writing - review & editing, project administration. **Allison Eriksson**: investigation, resources, data curation, writing - review & editing, project administration. **Cassia Man Ting Low**: investigation, writing - review & editing. **Josefine Waldthaler**: investigation, writing - review & editing. **Daniel Ferreira**: supervision, writing - review & editing. **Martin Ingvar**: conceptualization, writing - review & editing, resources, funding acquisition. **Per Svenningsson**: conceptualization, methodology, resources, supervision, data curation, writing - review & editing, Project administration, Funding acquisition. **Daniel Lundqvist**: conceptualization, methodology, formal analysis, resources, supervision, writing - original draft, writing - review & editing, project administration, funding acquisition.

