## Supplementary material for "Changes in non-oscillatory features of the cortical sensorimotor rhythm in Parkinson’s disease across age"

### 1 Model coefficients from regression analysis

#### 1.1 Regression coefficients for group, age, sex, and cortical thickness

Table S1: regression coefficients and 95% CI for the regression models of the sensorimotor signal features (Table 2) with Group, Age, Sex, and Cortical Thickness. Values in red indicate factors with Bayes Factor > 3 in the model comparison (see main text). LL: lower limit, UL: upper limit.

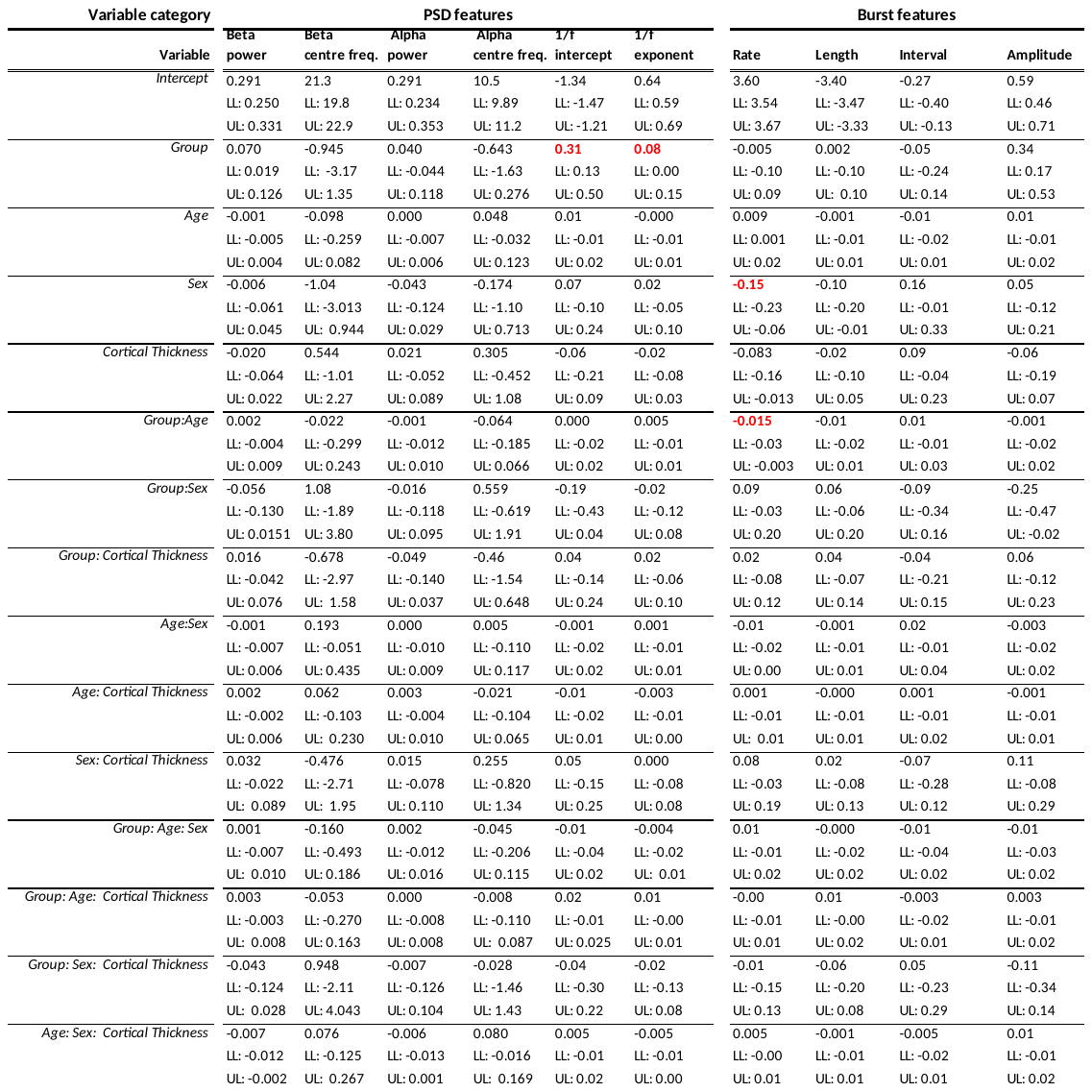

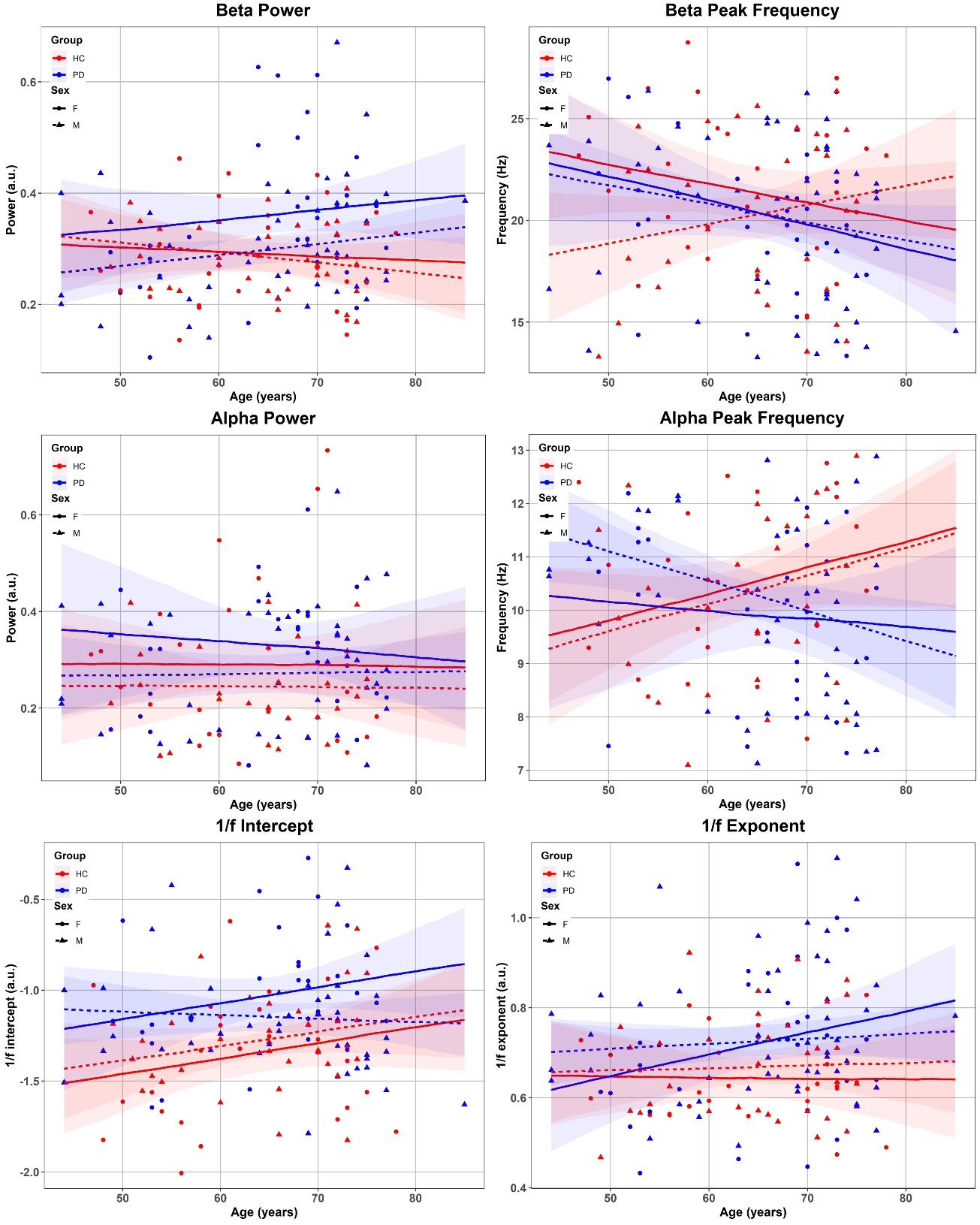

Figure S1: PSD feature over age for PD patients (blue) and healthy controls (red) split between females (solid lines) and males (dashed lines). The lines are the group-level predicted trends of the regression models. Shaded areas represent the 95% CI of the model prediction.

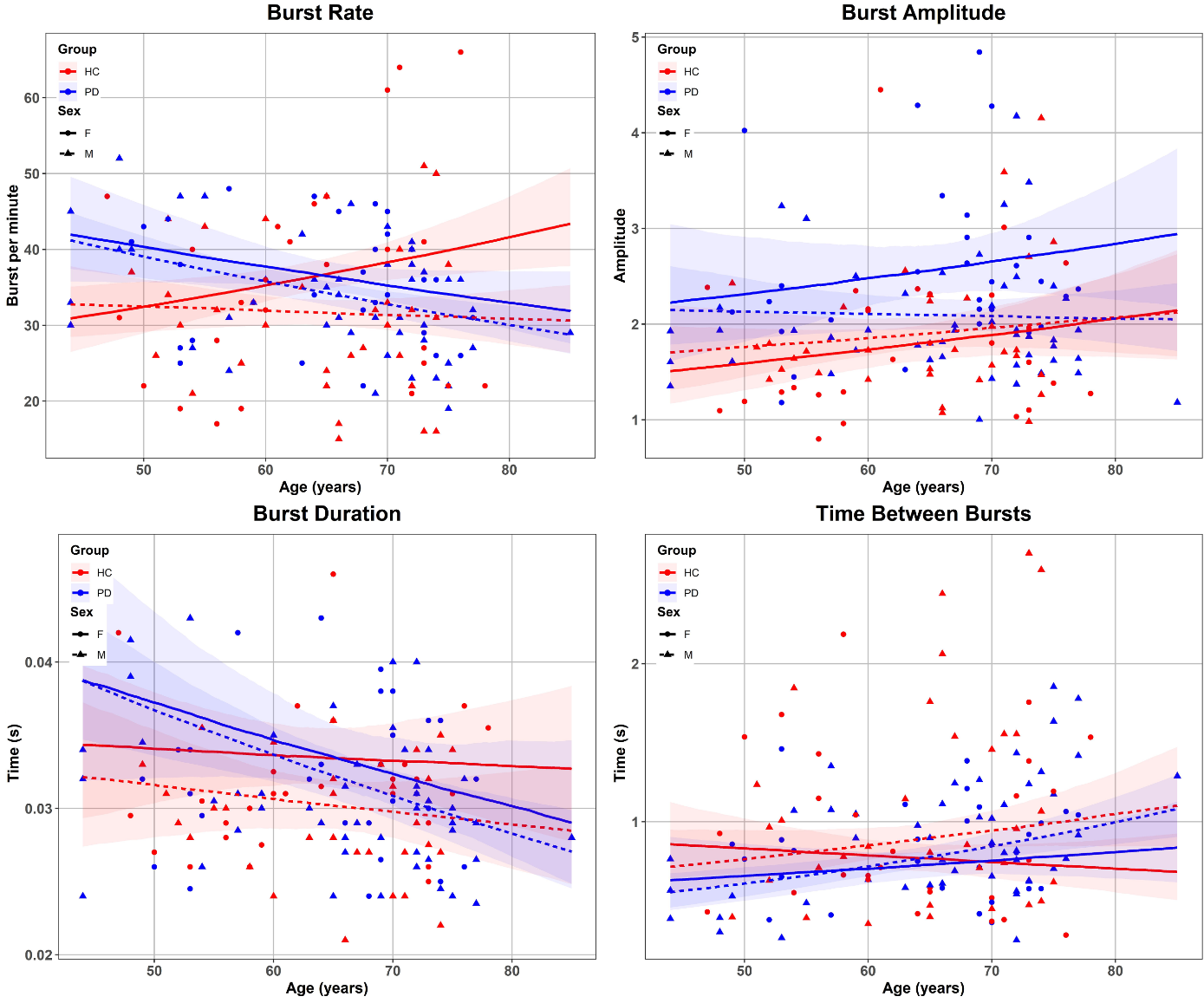

Figure S2: Burst features over age for PD patients (blue) and healthy controls (red) split between females (solid lines) and males (dashed lines). The lines are the group-level predicted trends of the regression models. Shaded areas represent the 95% CI of the model prediction.

#### 1.2 Regression coefficients for clinical scores

Table S2: Standardised regression coefficients (95% CI) for the six regression models on motor symptoms measured with the MDS-UPDRS-III. Values in bold indicate factors with Bayes Factor > 3 in the model comparison. LL: lower limit, UL: upper limit.

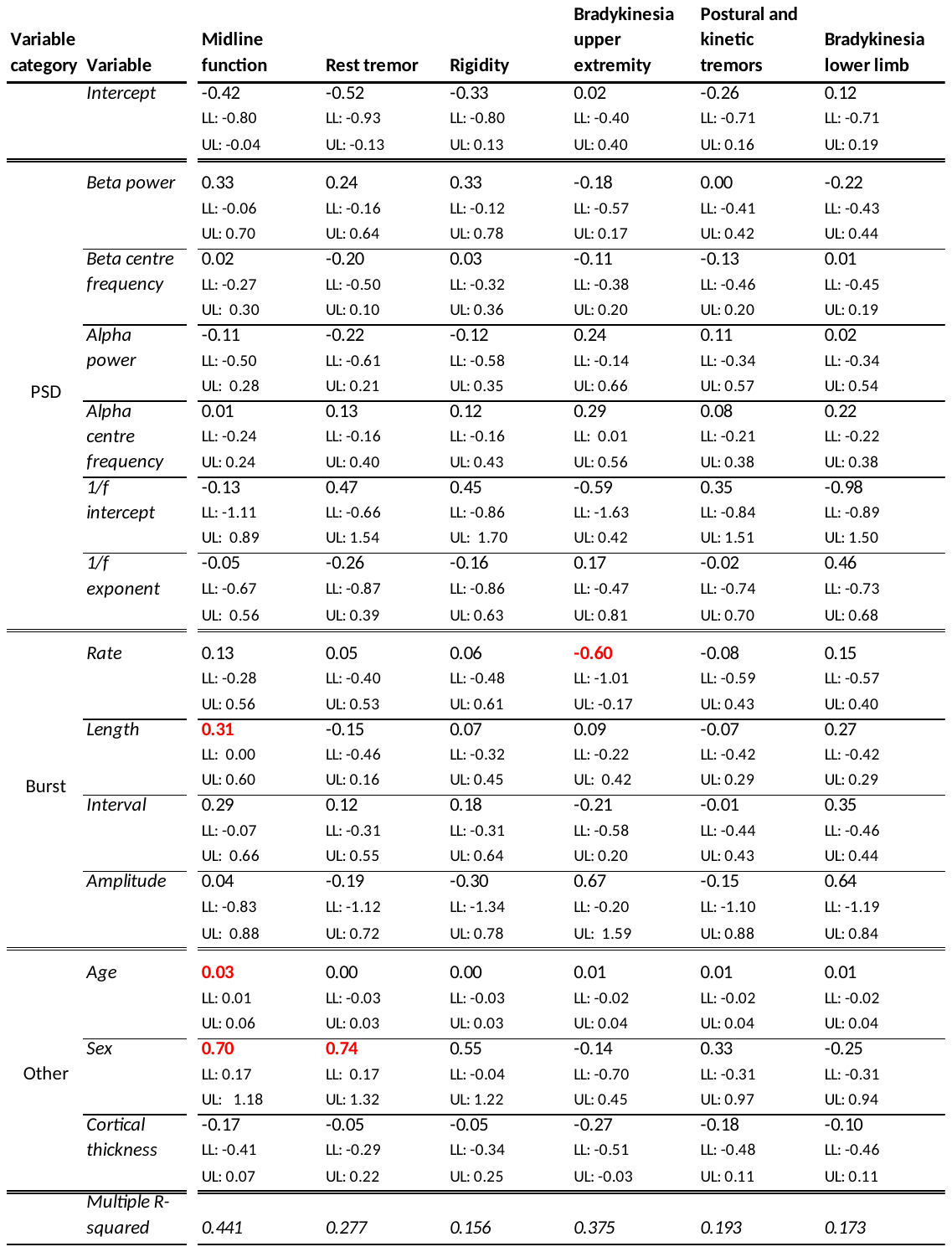

### 2  Bayes factors

Table S3: Bayes factors for the analysis of the effect of Group, Age, Sex, and Cortical Thickness on signal features (columns). BFs indicate comparisons for models with and without the factors in the rows in descending order. Red numbers indicate BF above the threshold (BF > 3) for substantial evidence for an effect. Blue numbers indicate BF below the threshold (BF < 1/3) for substantial evidence for no effect. Black numbers indicate BF in the inconclusive range (1/3 < BF < 3).

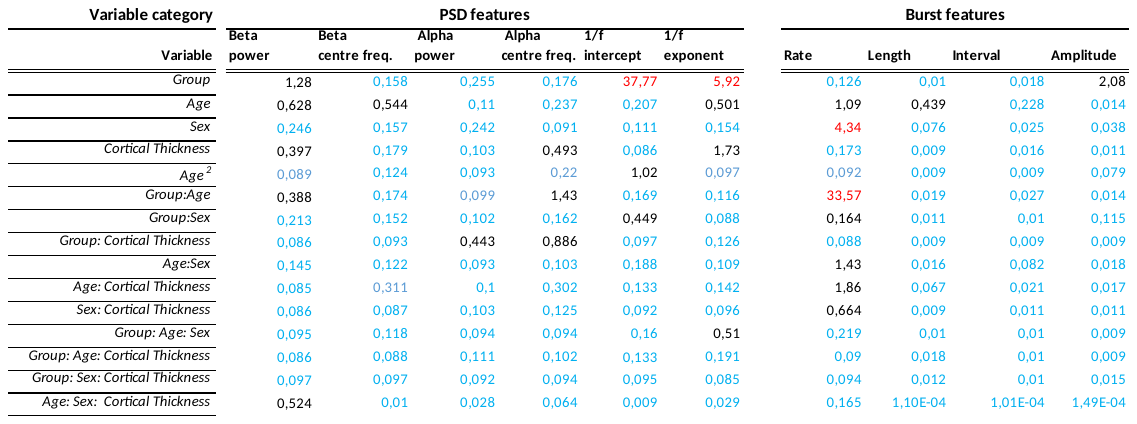

Table S4: Bayes factors for the analysis of signal features on clinical ratings of motor symptoms in the PD group. BFs indicate comparisons for models with and without the factors in the rows in descending order. Red numbers indicate BF above the threshold (BF > 3) for substantial evidence for an effect. Blue numbers indicate BF below the threshold (BF < 1/3) for substantial evidence for no effect. Black numbers indicate BF in the inconclusive range (1/3 < BF < 3).

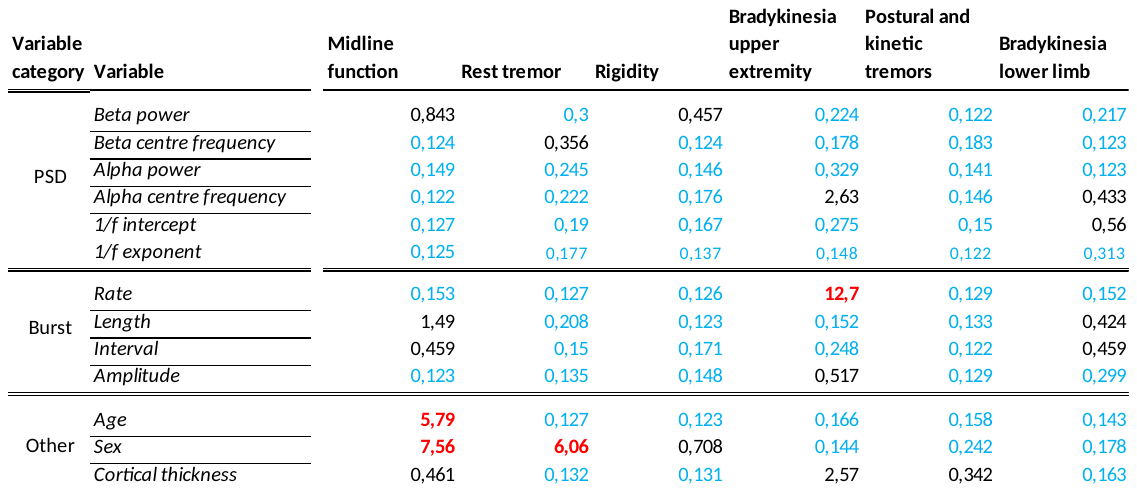

### 3 Correlation between variables

Table 5: Statistically significant (p < 0.05) correlation coefficients between all main variables. Green colours indicate a positive correlation between variables, and red colours indicate a negative correlation between variables—the intensity of the colour scales with the correlation coefficient.

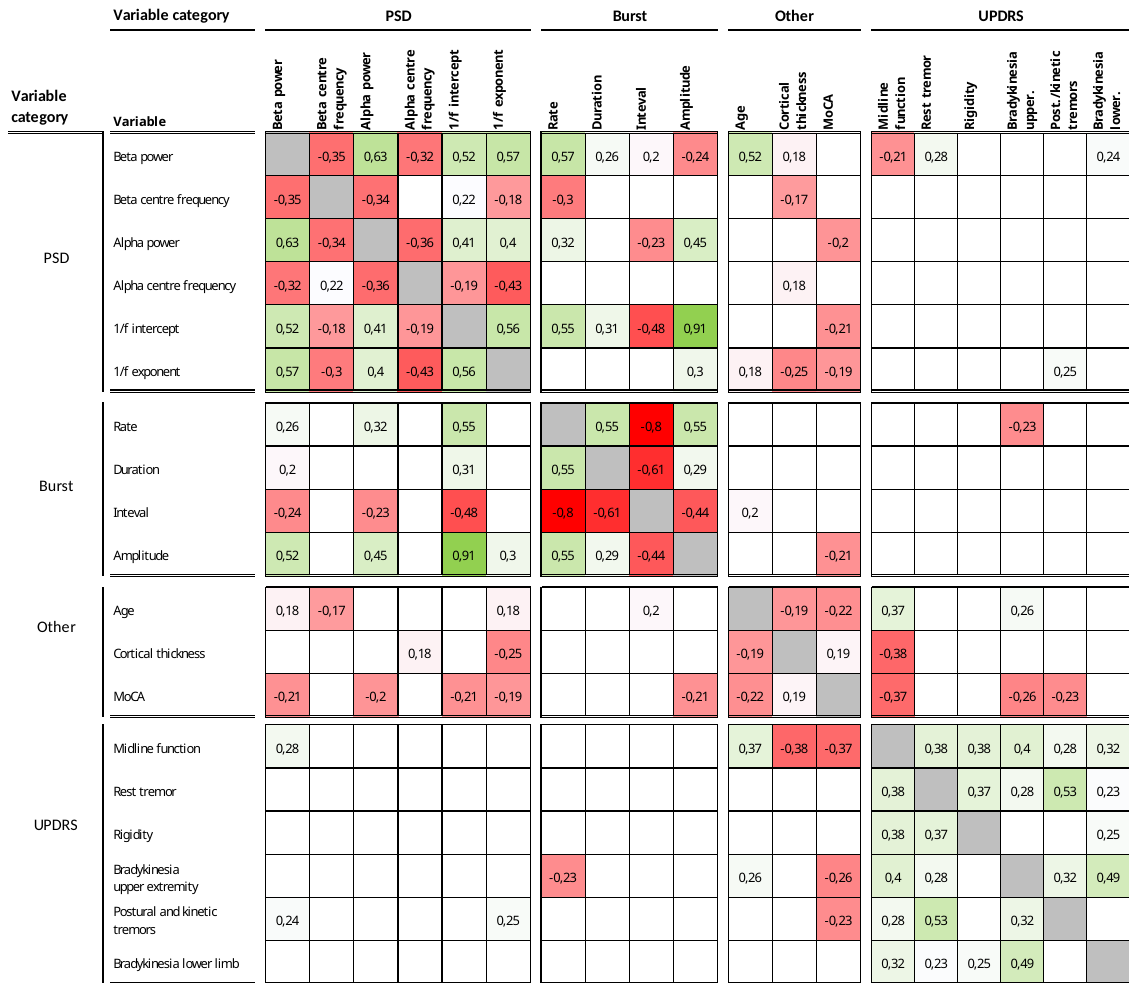
